## Supplementary Material for "Shifting patterns of dengue three years after Zika virus emergence in Brazil"

### Supplementary material for the manuscript "Shifting patterns of dengue three years after Zika virus emergence in Brazil."

**Text S1: Additional details on population initialisation**

Here we present an algorithm to initialise a synthetic population with the desired age distribution and serological profile. The central idea is to divide the procedure in two steps: first, we sample age and residual lifetime for each individual, and then generate an infection history that is compatible with serological data.

The first step consists in sampling age $A$ and residual lifetime $R$ from the joint distribution $p(A, R)$. Let us denote with $f(L)$ and $S(L) = \int_{L}^{\infty} f(x)dx$, respectively, the probability density and the inverse cumulative distribution of life-expectancy $L$. In order to sample from $p(A, R)$, we first sample $A$ from $S(A)/\bar{L}$, where $\bar{L}$ is the average life expectancy and then sample $L$ from $f(L)$, conditioned on $L$ being larger than $A$; finally, $R = L - A$ is the residual lifetime. This procedure guarantees that the population is already at demographic equilibrium at $t = 0$ where the age distribution is the one induced by the life-expectancy distribution $f(L)$.

The second step makes use of a sero-catalytic model to determine the number and timing of prior DENV infections, under the simplifying assumption that serotypes have spread independently in the past; more in detail, we assume that time τ since infection is distributed according to a non-homogeneous Poisson process with intensity $\lambda(x) = \lambda_1$ if $x \leq A_0$ and $\lambda(x) = \lambda_2$ if $x > A_0$. For each individual aged $A$ we then sample four infection times, one for each DENV serotype, and retain only those infection events such that $\tau < A$. Overall, this process implies that the probability of being seronegative at age $A$ is equal to $exp\left(-\ 4\int_{0}^{A} \lambda(x)dx\right)$.

Finally, we initialise the status of each individual at $t = 0$. If an individual is seronegative, it is automatically set to susceptible. Otherwise, we need to decide whether the host is incubating, infectious, cross-protected or susceptible after its most recent infection event, which we assume to have happened at time $\tau$ in the past due to serotype $i$. Now, let $t_E \sim Geom(\epsilon_D)$, $t_I \sim Geom(\sigma_D)$ and $t_R \sim Geom(l_D)$: if $t_E > \tau$ we set the individual exposed to $i$; if not, we check if $t_E + t_I > \tau$, in which case we set the individual infectious with $i$. If not, we check if $t_E + t_I + t_R > \tau$, in which case we set the individual as cross-protected, after having recovered from $i$. If this is still not the case, we set the individual as susceptible (on top of being fully immune to $i$).

Alternatively, it is possible to initialise a population's serological profile in a purely mechanistic way, by letting the population be exposed to DENV serotypes for a long enough period. More in detail, we initialise the age profile as before, but let DENV spread for 100 years before introducing ZIKV and/or collecting any other information. Furthermore, at $t = 0$ we randomly select a proportion $1 - R_{0,D}^{-1}$ of hosts for each DENV serotype and immunise them to that serotype.

### Supplementary Figures

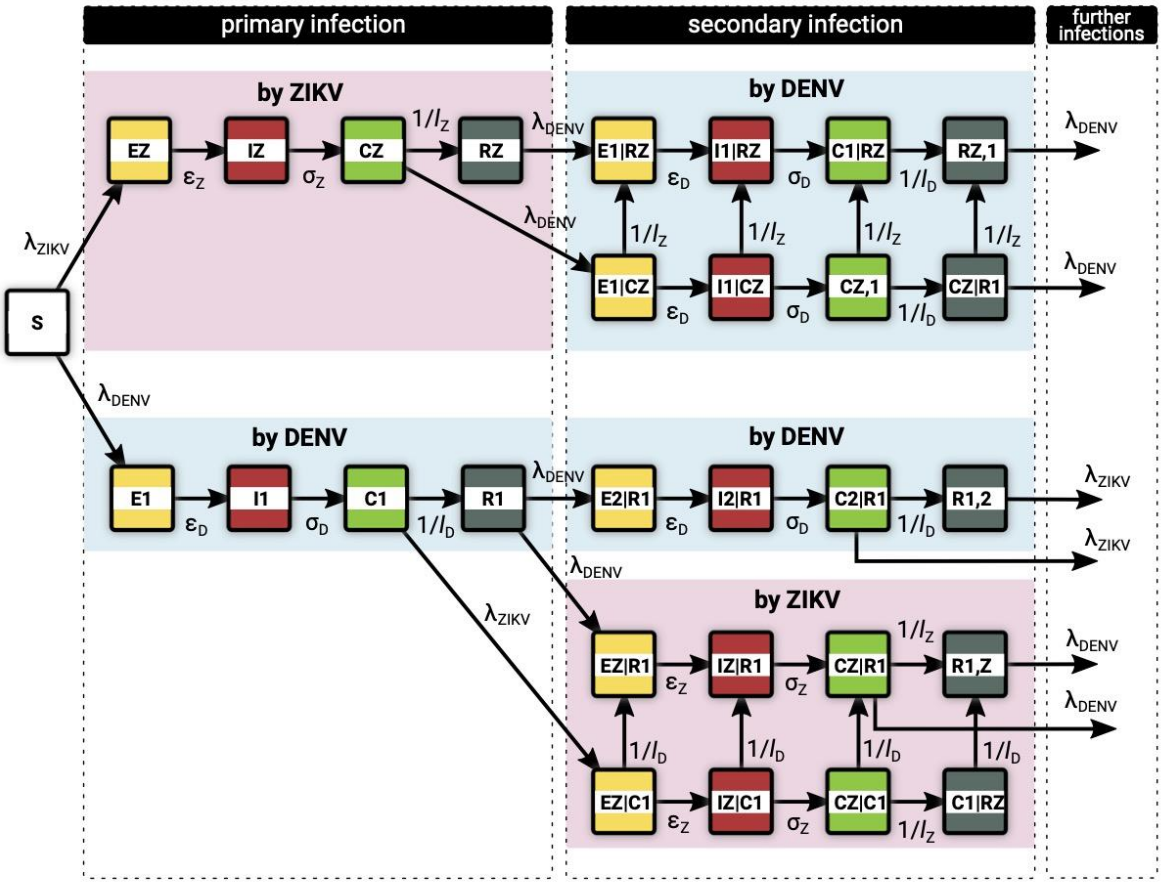


**Figure S1. Model diagram.** Each individual is born susceptible (S) and can be infected with ZIKV or any DENV serotype at rates $\lambda_{ZIKV}$ and $\lambda_{DENV}$, respectively (primary infection). The force of infection of each pathogen is proportional to its prevalence and can be affected by seasonality. Following infection with ZIKV (DENV), an individual enters first a latent compartment (E) and then becomes infectious (I) at rate $\epsilon_Z$ ($\epsilon_D$). While infectious, recovery occurs at rate $\sigma_Z$ ($\sigma_D$). Recovered individuals first enter a cross-protected compartment (C). Cross-protection is transient and is lost at rate $l_Z^{-1}$($l_D^{-1}$). After that, the host only maintains immunity against reinfection with the same serotype (R). Cross-protection induced by ZIKV (DENV) prevents onward transmission of DENV (ZIKV), but does not affect susceptibility to infection. In addition, DENV-induced cross-protection prevents infections with distinct DENV serotypes. Note that co-infection is not possible in this model.

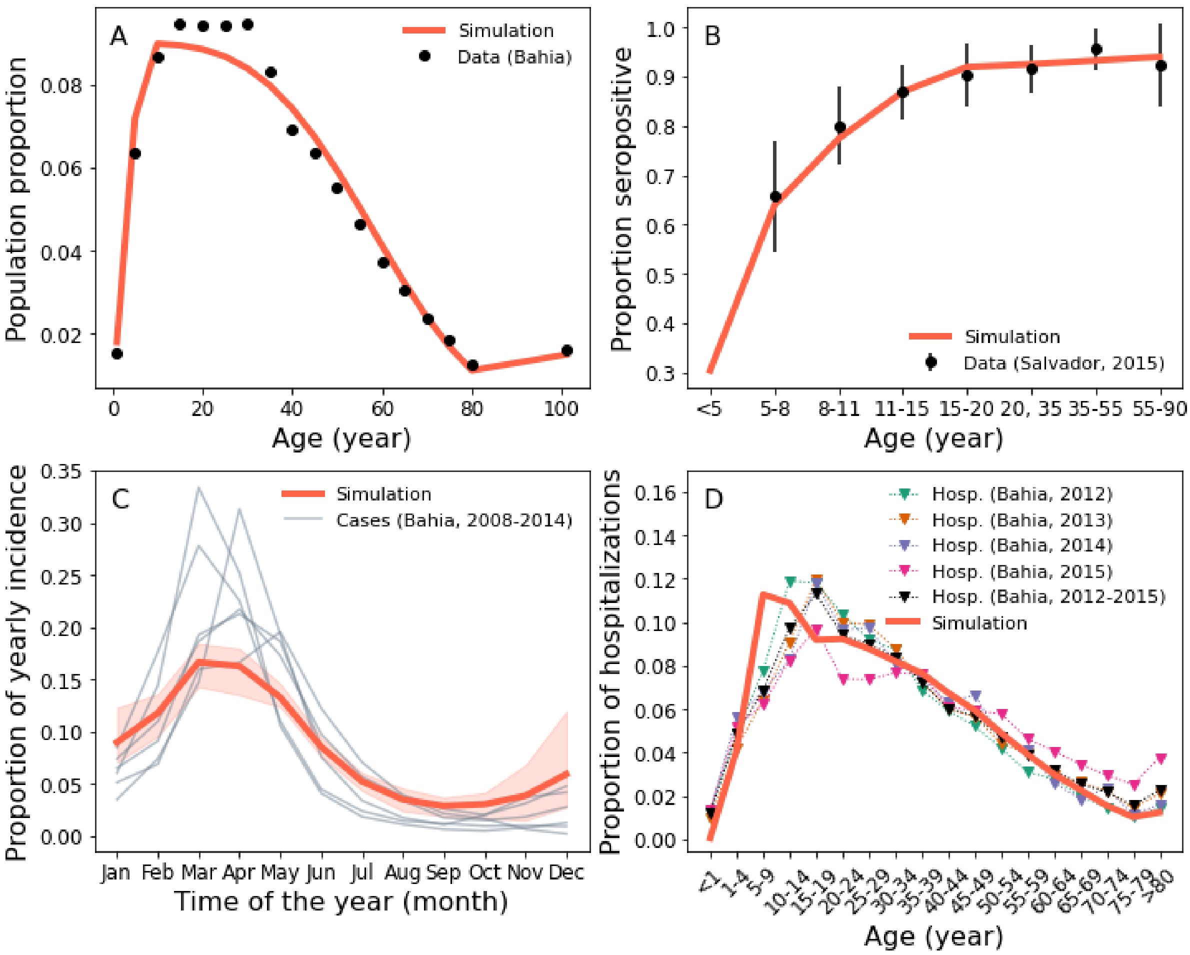


**Figure S2. Calibrating host population and DENV endemic dynamics.** (A) Simulated (line) and empirical (dots) population age distributions. The latter was obtained by averaging over Bahia age distributions from 2008 to 2018. (B) Simulated (line) vs observed (dots) seroprevalence profiles. The simulated seroprevalence profile is based on a non-homogeneous Poisson process with intensities $\lambda_1 = 0.04\, y^{-1}$, $\lambda_2 = 0.0014\, y^{-1}$ and a breakpoint $A_0 = 16y$. Simulated and empirical profiles are based on findings in (Rodriguez-Barraquer et al. 2019). Bars represent the 95% C.I., based on a binomial distribution assumption. (C) Simulated cases (red) vs observed incident Dengue hospitalizations (gray, notified DENV cases) in Bahia from 2008 to 2014. (D) Age distribution of secondary DENV infections in simulations (red line) and observed hospitalizations in Bahia from 2012 to 2015 (markers). In C and D incidence is obtained from the fourth and the first year since the beginning of simulations, respectively.

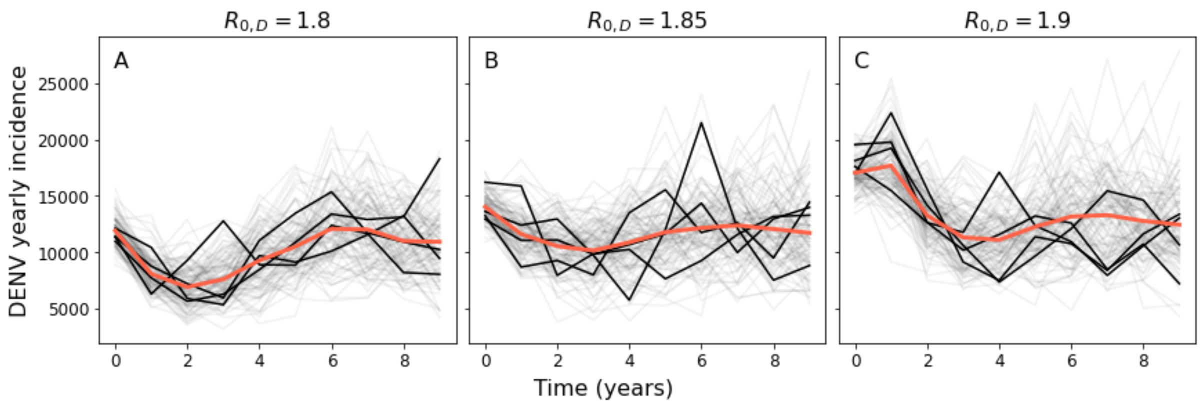

**Figure S3. Calibrating $R_{0,D}$.** Panels show DENV yearly incidence from 100 simulations (dim lines) for increasing values of $R_{0,D}$. A few simulations are highlighted in black, while the mean trend is shown in red. Here, the host population and DENV parameters are set as in Fig. S6. In particular, individual immune histories are initialised according to the scheme outlined in Text S4. At endemicity, one would expect yearly incidence to fluctuate around a stationary baseline value, as recovered in the middle panel. Selecting a too small or too large value of $R_{0,D}$ would instead introduce some 'distortion' in DENV dynamics (A,C) because of a mismatch between $R_{0,D}$ and the initial level of immunity implied by the imposed initial serological profile. Please note that it is not possible to recover the exact serological profile in Fig. S6B by simply simulating DENV dynamics using a fixed $R_{0,D}$. Nonetheless, this is not a major concern in our analysis since we are only interested in a short time horizon after ZIKV introduction.

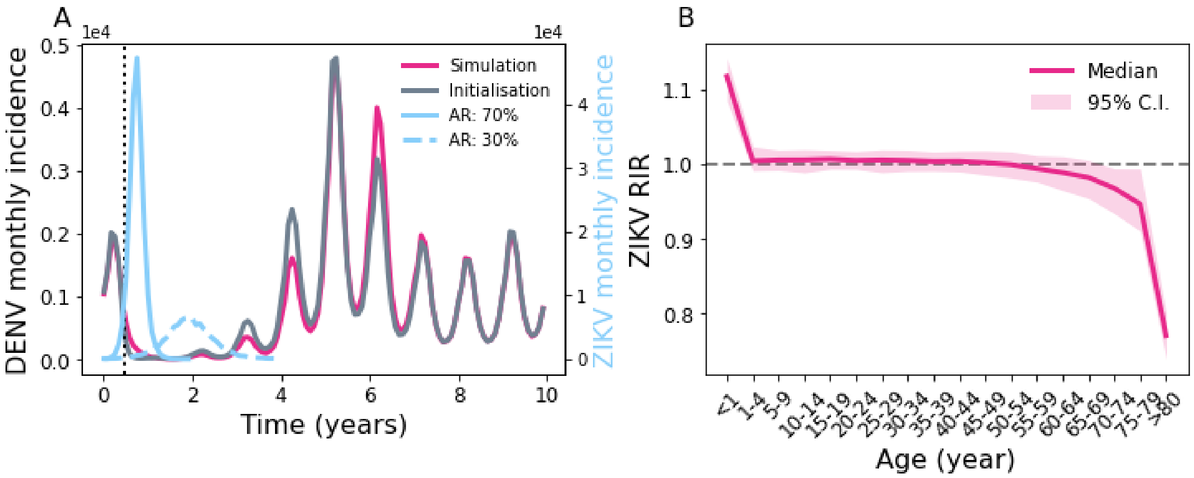


**Figure S4. Comparison of ZIKV seeding methods.** (A) Median monthly DENV (grey or fuchsia) incidence over 10 years. With the serology-aware initialisation method (grey line, see also Text S4), a proportion $\rho_Z = 0.7$ of the population becomes suddenly infected with ZIKV at $t = 0.45$ years (vertical line), but onward viral transmission is not allowed. This is compared with another simulation method where ZIKV infects roughly the same amount of the population starting from just 10 infectious seeds selected at $t = 0$ (fuchsia). Here, the desired attack rate is obtained by inverting the final epidemic size formula for a SIR model, namely $R_{0,Z} = -log(1 - \rho_Z)/\rho_Z$ (Miller 2012), and assuming no seasonality in ZIKV transmission ($\beta_{1,Z} = 0$). The same panel also shows ZIKV incidence for two simulations where 70% and 30% of the population becomes infected (solid and dashed blue lines, respectively). Note that the second epidemic is slower and unfolds over multiple years, highlighting technical problems encountered in simulating epidemics with low $R_{0,D}$. (B) ZIKV Relative Infection Risk (median and 95% IQR) using the simulation method to seed ZIKV. The flat profile implies a uniform attack rate over most age classes, which is also implied by the serology-aware initialisation method by construction. In both scenarios, $l_Z = 1$ y. DENV is allowed to spread for a burn-in period of 100 years before the period shown in the plot and $R_{0,D} = 1.65$. Other parameters are as in the default scenario. Results are averaged over 200 simulations.

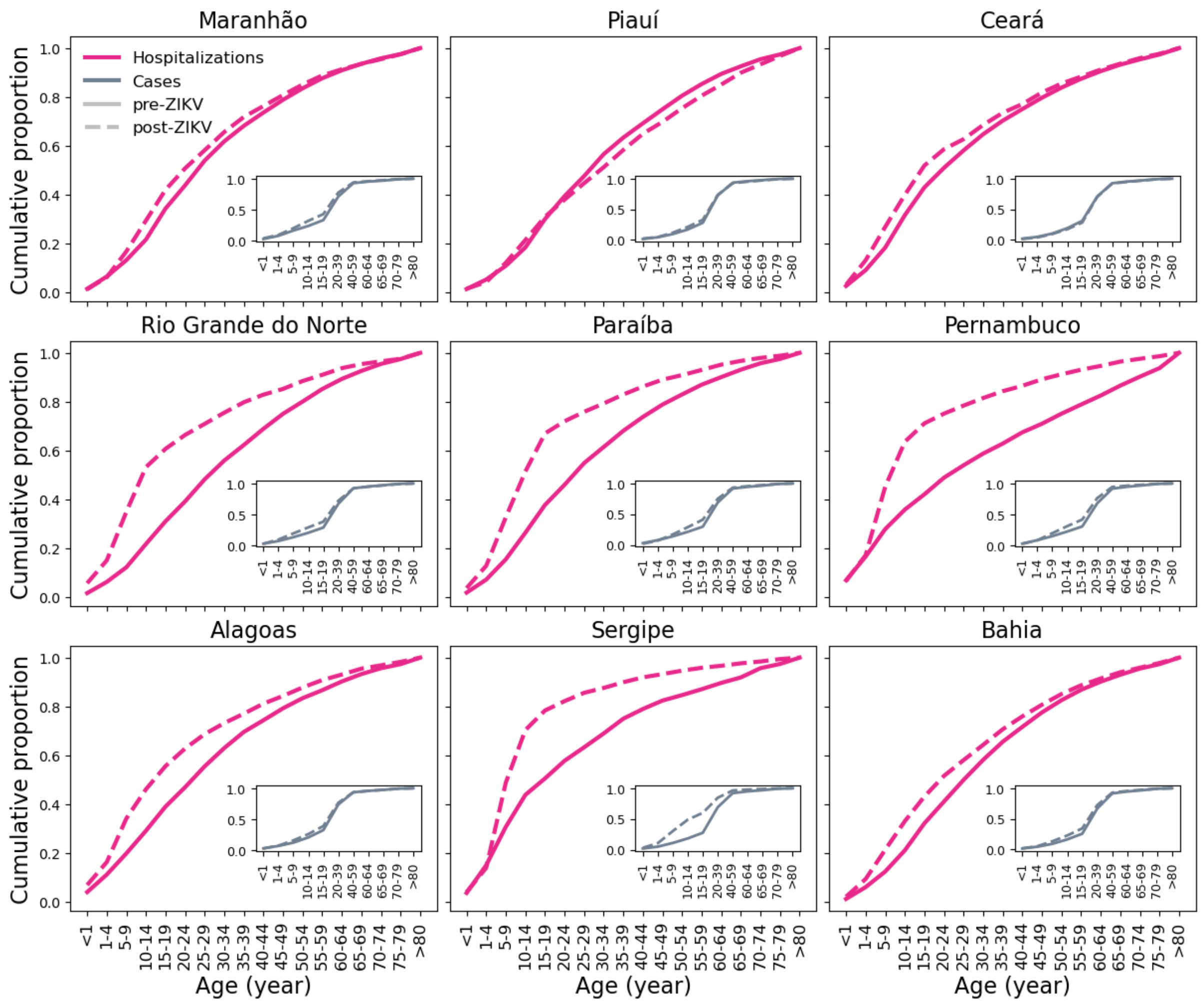


**Figure S5. Age distribution of DENV in the North-Eastern region of Brazil.** Cumulative age distributions for hospitalizations (fuchsia, main panel), and cases (grey, inset) before and after ZIKV (solid and dashed lines respectively). Each panel denotes a different state in the North-East. Pre- and post-ZIKV distributions are based on data from the periods 2013-2015 and 2018-2019, respectively.

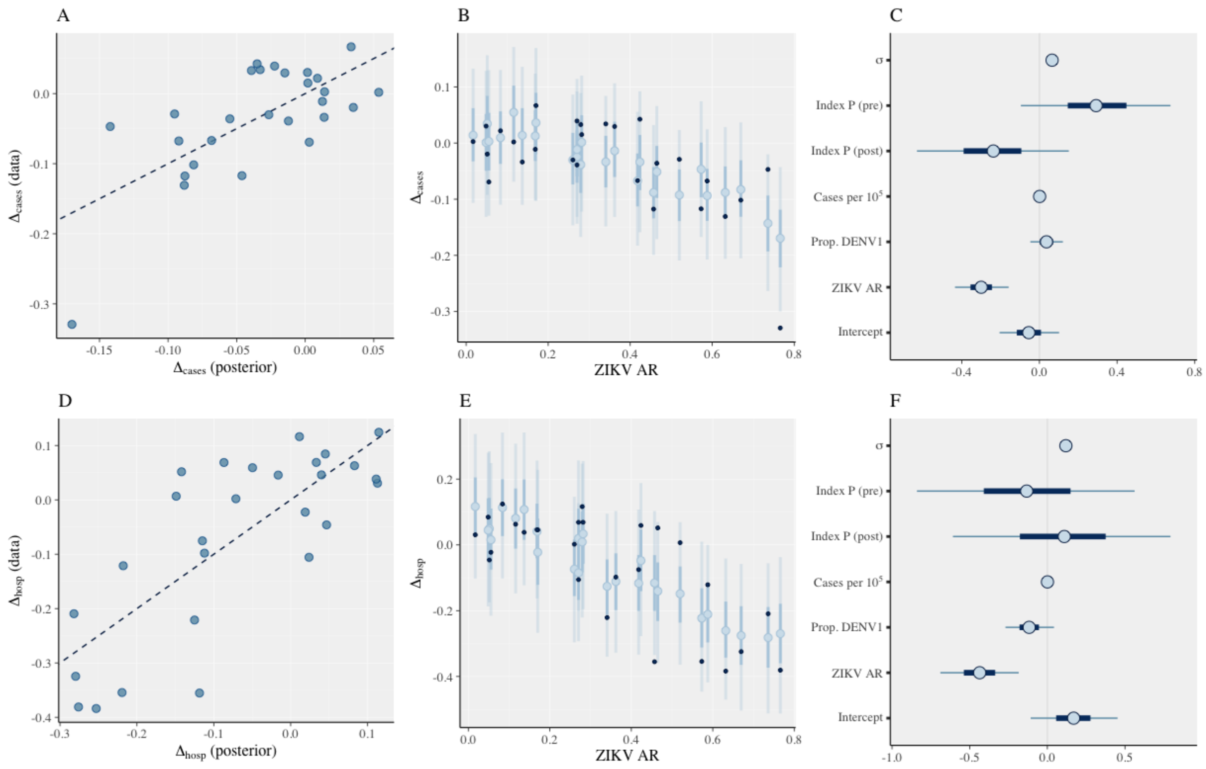


**Figure S6. Linear regression analysis.** (A) Observed vs fitted values. The line is a guide for the eye (B) Posterior median (blue circles) and central intervals estimates (50% and 90%) for $\Delta_{cases}$ with observed data overlaid (black dots), plotted against ZIKV attack rate. (C) Posterior median and central intervals (50% and 90%) estimates for regression coefficients, intercept term and error standard deviation ($\sigma$). (D-F) Same as above, but with $\Delta_{hosp}$ as the dependent variable. The regression coefficient relative to estimated ZIKV attack rate is significantly different from 0 for both case and hospitalisation data. Furthermore, ignoring ZIKV attack rate had a significantly negative effect on predictive accuracy, while adding region-specific intercepts did not improve model fit noticeably (not shown). Results are based on 4000 posterior samples collected from 4 independent chains after an initial warmup of 1000 steps.

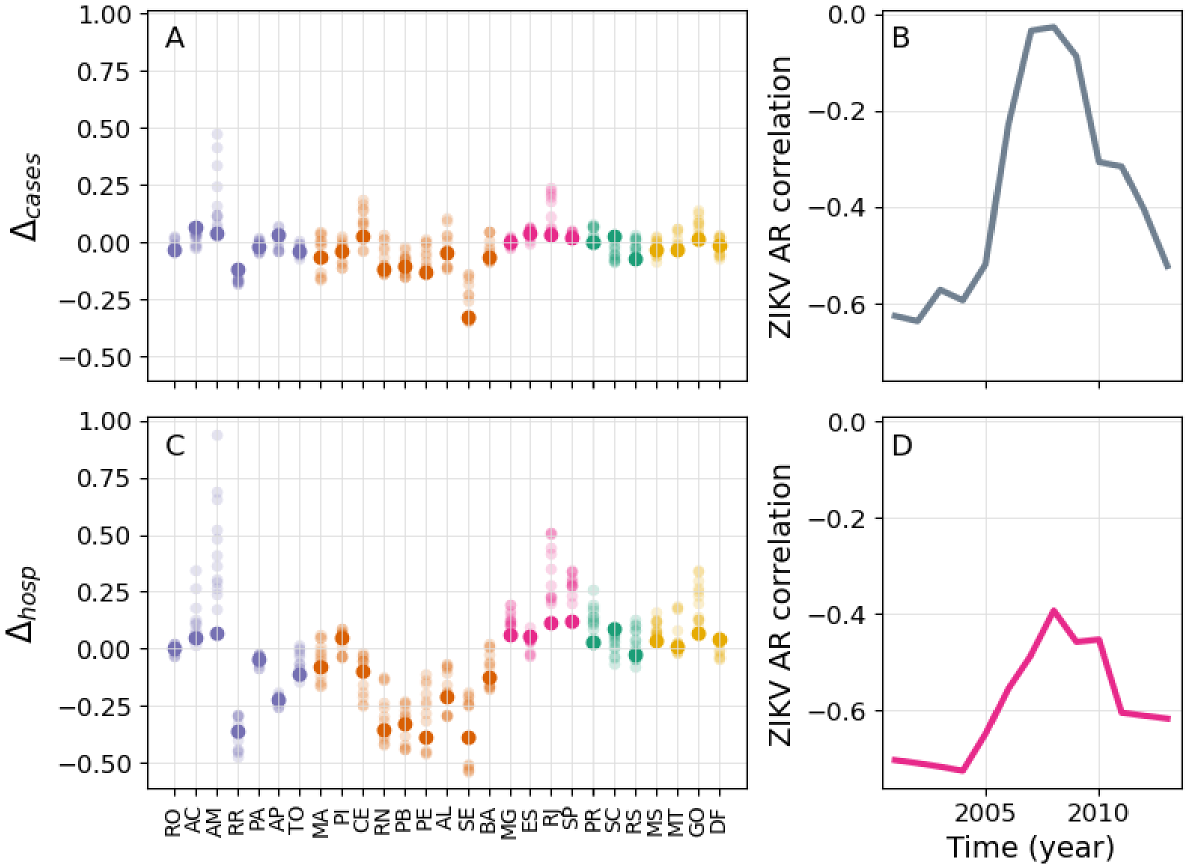


**Figure S7. Age variation with respect to different time periods.** (A) We calculate the age shift $\Delta = (\bar{A}_{post} - \bar{A}_{pre})/\bar{A}_{pre}$ for different states and with respect to different reference periods to estimate $\bar{A}_{pre}$; for each state, $\bar{A}_{post}$ is fixed and calculated based on cases reported in the period 2018-2019. Solid circles denote the latest estimate based on the period 2013-2015, while dim circles correspond to earlier periods. (B) Spearman rank correlation coefficient between $\Delta$ and ZIKV attack rate using different reference periods to estimate $\bar{A}_{pre}$, indexed by the starting year $t$; the period itself ends in year $t + 2$. (C,D) Same as panels (A,B), but using hospitalisation data instead of reported cases. Panels suggest that the statistical association between ZIKV attack rate and $\Delta$ is robust with respect to the choice of the reference time frame, although the statistical signal is somewhat diminished when the pre-ZIKV period includes years between 2007 and 2010. It should be noted, however, that two serotype changes occurred in 2007 and in 2009, due to DENV2 and DENV1 respectively. These events may partially explain the observed loss in statistical signal.

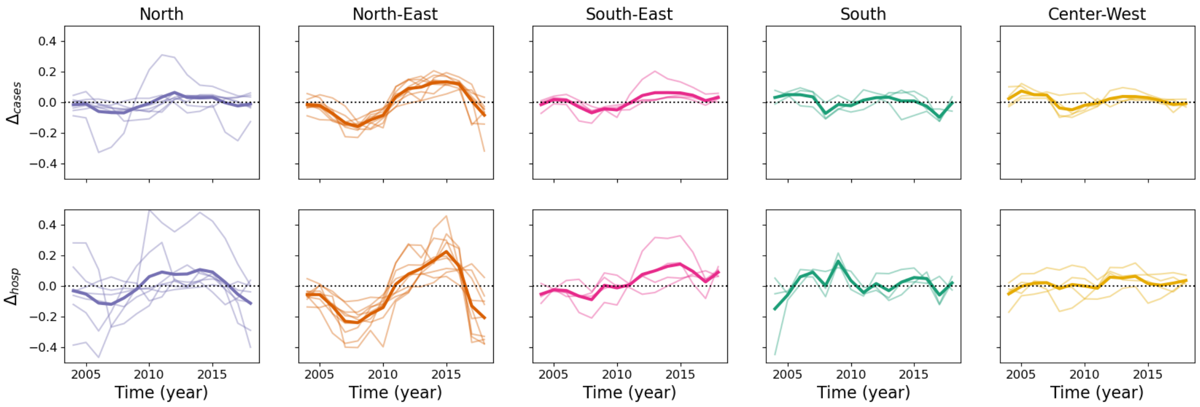

**Figure S8. Dynamics of $\Delta$ over time.** Panels show $\Delta$ for both reported cases (top row) and hospitalizations (bottom row). For each year $t$ we compute $\Delta(t) = (\bar{A}_{post} - \bar{A}_{pre})/\bar{A}_{pre}$, where $\bar{A}_{post}$ aggregates incidence from years $t$ to $t + 1$, and $\bar{A}_{pre}$ aggregates incidence from year $t - 5$ to $t - 3$ . For example, $\Delta(2018)$ compares incidence in 2018 and 2019 with incidence from 2013 to 2015. Thick lines represent the average over all states in the same region, while individual states are displayed as dim lines.

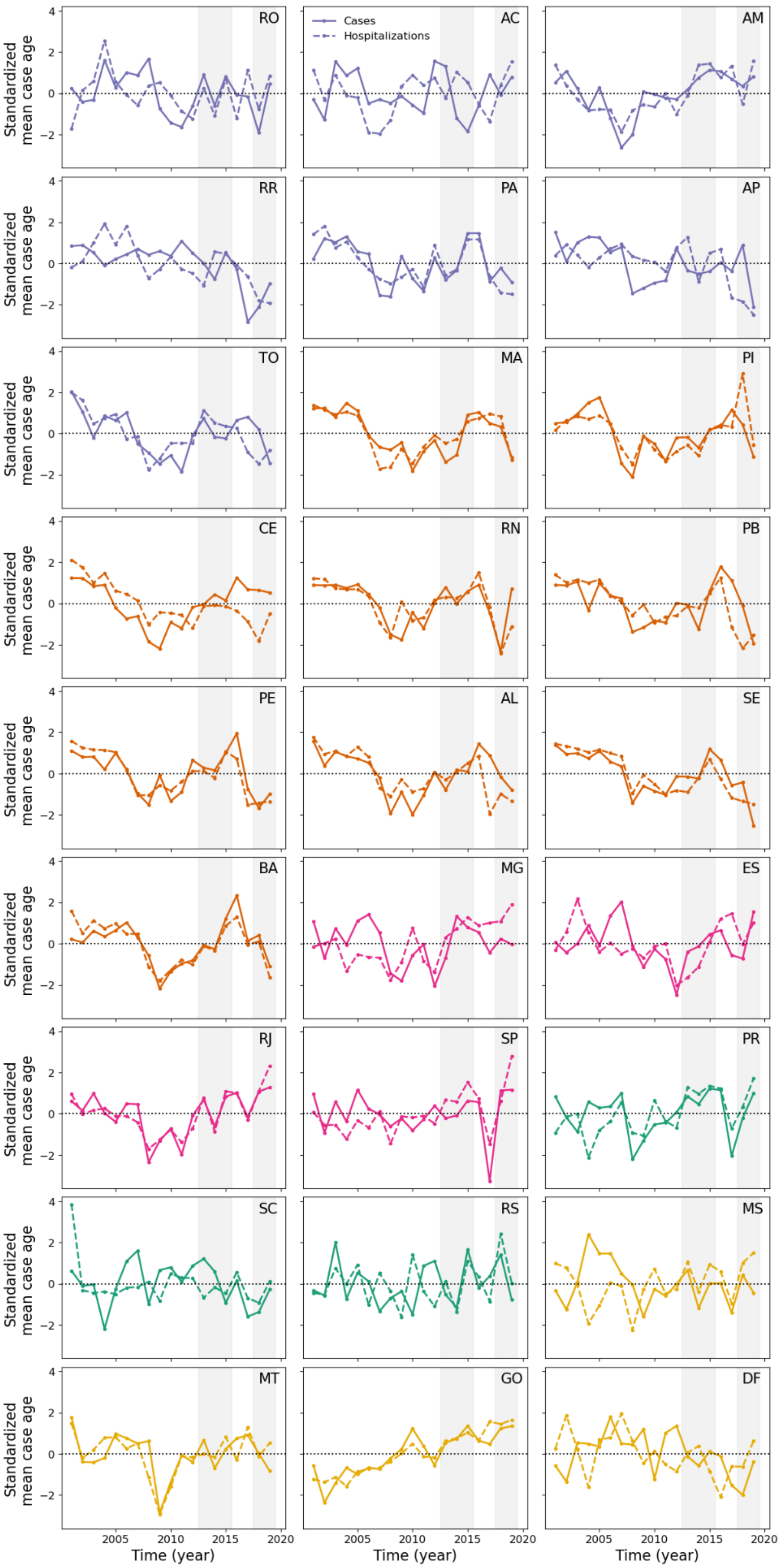

RO
AC
AM
RR
PA
AP
TO
MA
PI
CE
RN
PB
PE
AL
SE
BA
MG
ES
RJ
SP
PR
SC
RS
MS
MT
GO
DF
Cases
Hospitalizations
Standardized mean case age
Time (year)
4
2
0
−2
2005
2010
2015
2020

**Figure S9. Evolution of $\bar{A}$ over time across Brazil.** Each panel displays $\bar{A}$ for notified DENV cases (solid) and hospitalizations (dashed) from 2001 to 2019 for a single state. $\bar{A}$ was standardised by subtracting its mean value and scaled by its standard deviation computed over the same period. Colour code is the same as in Fig. 3 in the main manuscript. Grey shaded areas correspond to pre- and post-ZIKV periods considered in the main analysis.

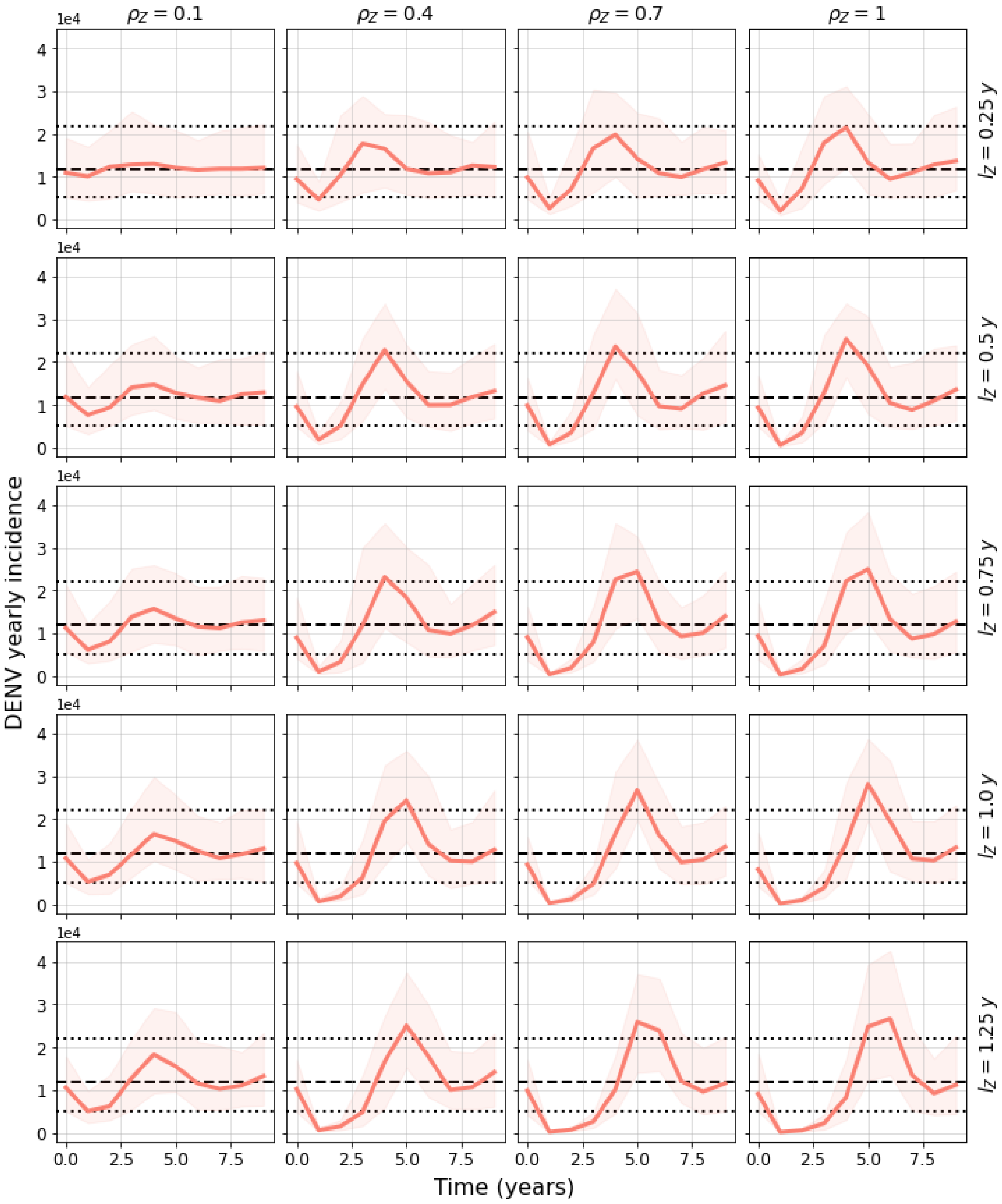


**Figure S10. Impact of ZIKV on DENV resurgence.** Each panel shows median DENV yearly incidence (solid red line) over 10 years, ZIKV being introduced at $t = 0.45$ y, for different ZIKV attack rates (increasing from left to right) and duration of ZIKV-induced cross-protection (increasing from top to bottom). The shaded area represents the 95% incidence interquantile range computed from 200 simulations. Simulations show that temporary cross-immunity induced by ZIKV can drive DENV incidence to abnormally low levels with respect to a scenario in which ZIKV never occurred. Median and 95% interquantile range DENV incidence in such baseline scenarios are shown as dashed and dotted lines, respectively. DENV circulates at low levels from 2 to 3 years depending on ZIKV attack rate ($\rho_Z$) and duration of cross-protection ($l_Z$), in agreement with a previous simulation study (Borchering et al. 2019). To obtain these plots, we first allow DENV to spread for a burn-in period of 100 prior to $t = 0$. ZIKV is introduced by infecting a proportion $\rho_Z$ of hosts, as detailed in the main text. Here, $R_{0,D} = 1.65$. Other parameters are set to default values.

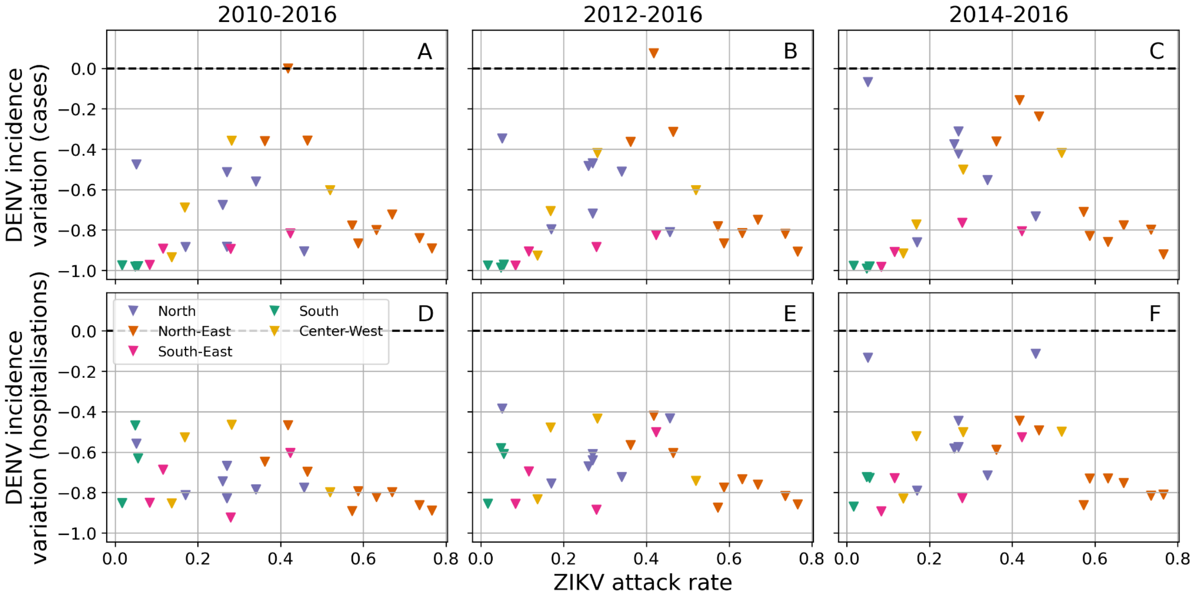


**Figure S11. Reduction in DENV incidence following the emergence of ZIKV.** In each panel, scatters represent state-specific relative variation in mean DENV yearly incidence before and after ZIKV against estimated ZIKV attack rate. First and second rows are based on reported cases and hospitalisations, respectively. More in detail, we compare DENV incidence in 2017 with incidence from three different periods, namely 2010-2016 (A,D), 2012-2016 (B,E) and 2014-2016 (C,F). Almost all states experienced a reduction in both DENV reported cases and hospitalisations in 2017 with respect to previous years, although no firm conclusion on the relationship between DENV reduction and ZIKV attack rate can be drawn based on these data. Interestingly, DENV transmission was particularly disrupted in states with either low or high ZIKV attack rates. It should be noted, however, that historical trends of DENV circulation may vary considerably from state to state (Grubaugh, n.d.), potentially confounding our analysis.

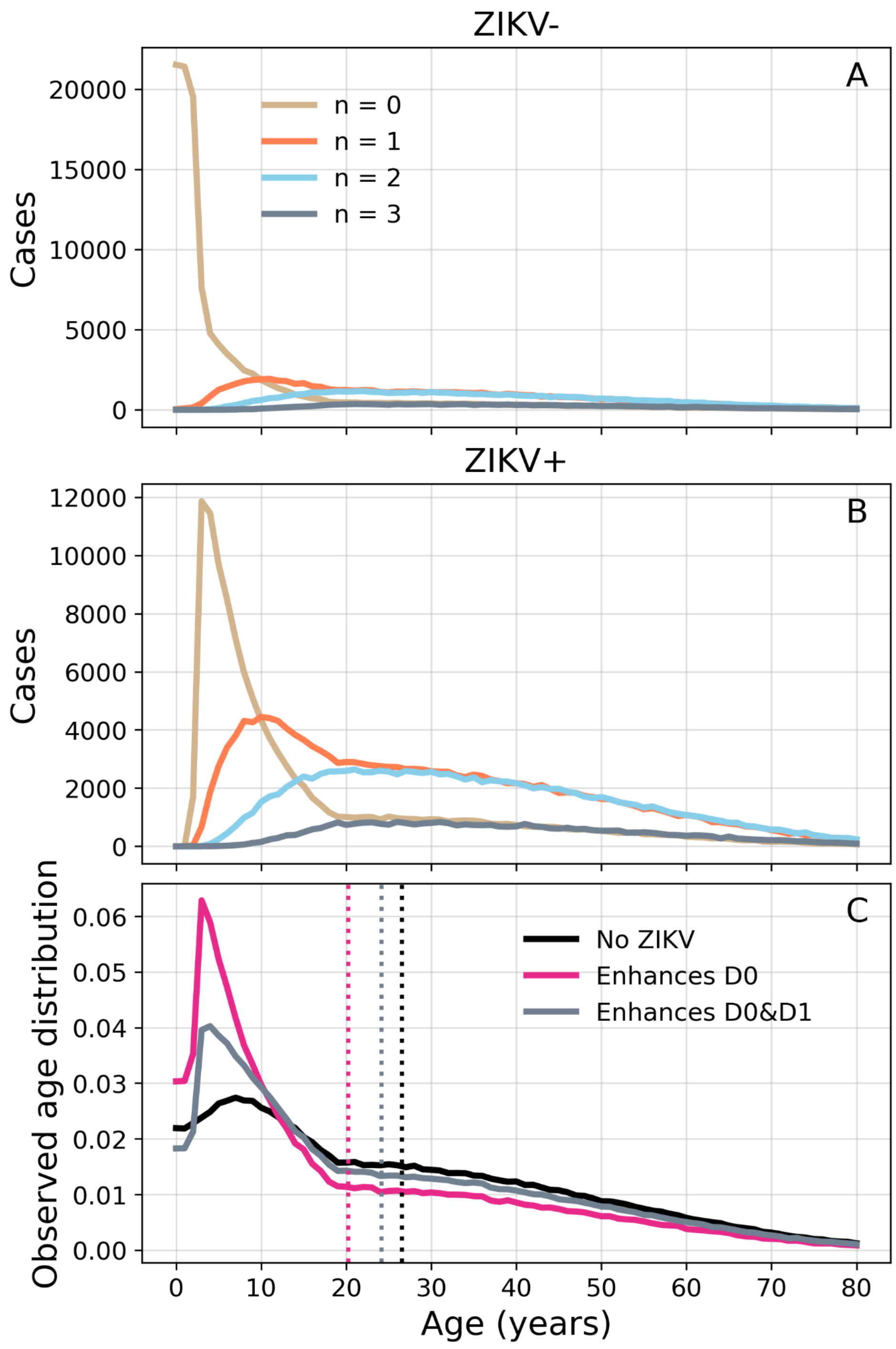


**Figure S12. DENV seroincidence.** Counts of incident DENV cases by number $n = 0, 1, 2, 3$ of past DENV infections in ZIKV- (A) and ZIKV+ (B) hosts, assuming a 70% ZIKV attack rate. Case counts are collected during the fourth year since simulation start time and aggregated over multiple simulations. (C) Distribution of observed incident DENV cases, assuming that ZIKV enhances severity in primary DENV infections only (fuchsia, mechanism (1) in the main manuscript) and both primary and secondary infections (dark grey, mechanism (2) in the main manuscript). Dotted lines represent the mean observed case age from the corresponding distribution. (C) demonstrates that mechanism (1) is able to generate a larger decrease in observed case age than mechanism (2), with respect to a reference scenario with no ZIKV (black). Host population and transmission parameters are set to default values. The underlying explanation is that secondary DENV infections in ZIKV+ hosts (orange lines in (A,B)), whose severity is enhanced by mechanism (2) but not (1), typically affect older hosts than primary DENV infections (beige lines). Overall, both mechanisms can lower the mean observed case age.

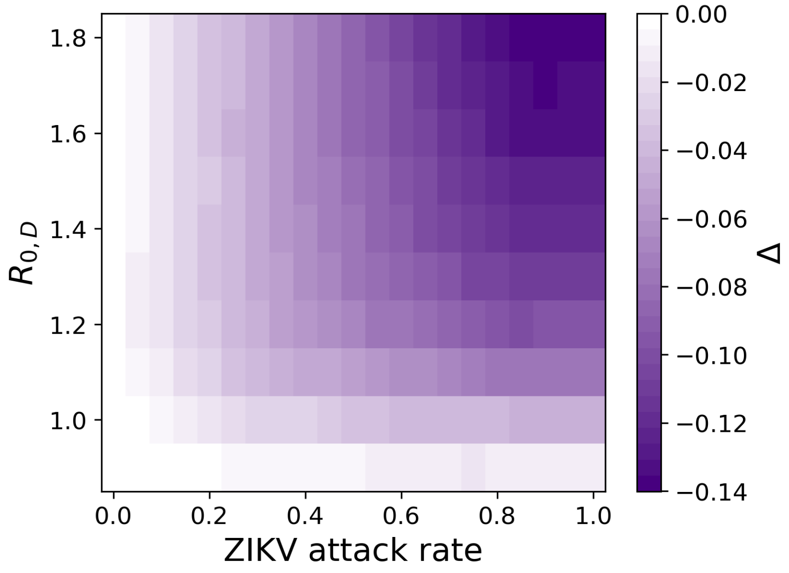


**Figure S13. Mean age shift under different ZIKV and DENV transmission scenarios.** The heatmap mirrors Fig. 4C in the main text under the assumption that ZIKV enhances both primary and secondary DENV infections (mechanism (2) in the main text). Incidence weights $w_n^{-/+}$ are the same as those specified in Fig. 4 in the main manuscript for mechanism (2). Δ is calculated using incidence from the fourth year since simulation start time. DENV is allowed to spread for a burn-in period of 100 years before ZIKV is introduced. We average results over 300 independent realisations.

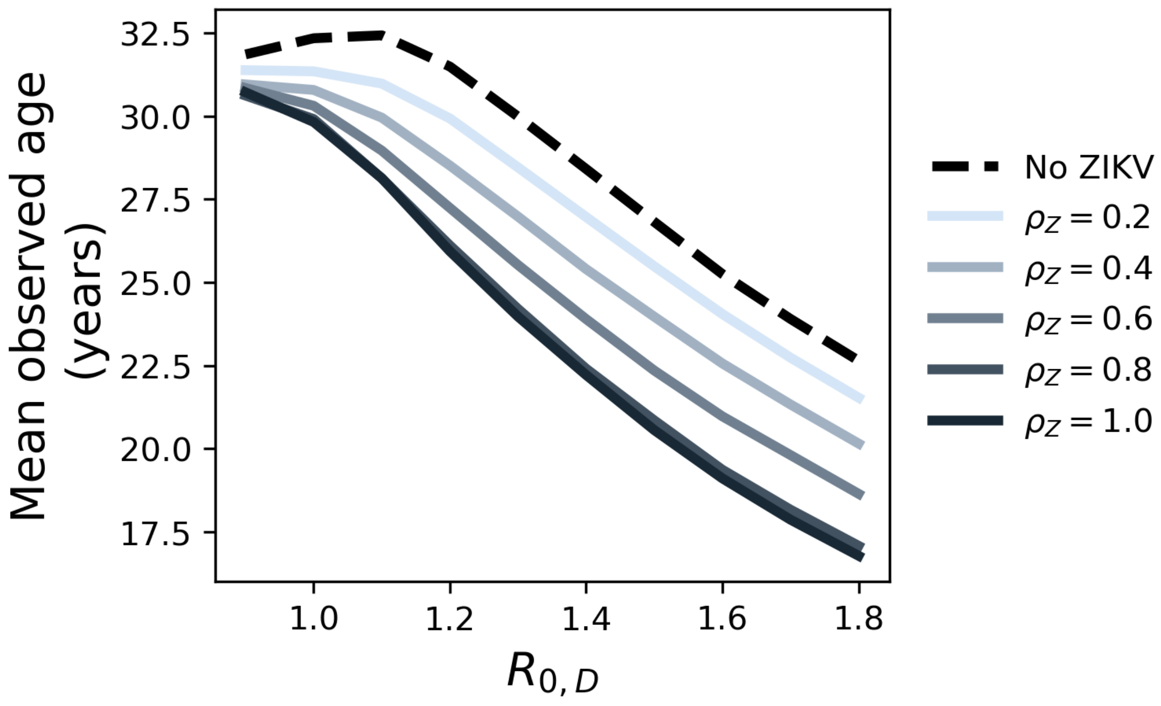


**Figure S14. Impact of baseline DENV transmission.** Mean observed age of incident cases as a function of $R_{0,D}$ and for different values of ZIKV attack rate ($\bar{A}_{post}$). The dashed line corresponds to simulations without ZIKV and is thus equivalent to $\bar{A}_{pre}$, the mean observed case age before ZIKV is introduced. Results are based on the assumption that ZIKV enhances disease in primary DENV infections only (mechanism (1) in the main text).

The mean observed case age before ($\bar{A}_{pre}$) and after ($\bar{A}_{post}$) ZIKV, which enter Δ's definition through $\Delta = \bar{A}_{post}/\bar{A}_{pre} - 1$, do not depend on $R_{0,D}$ in the same way. Typically, $\bar{A}_{post}$ decreases with $R_{0,D}$ (solid lines), while $\bar{A}_{pre}$ initially increases with $R_{0,D}$ and then declines (dashed line). Taken together, these results mean that Δ must decrease more rapidly at low $R_{0,D}$. To obtain these plots, we let DENV spread for a burn-in period of 100 years before ZIKV is introduced at $t = 0.45$ y. We compute DENV incidence during the fourth year since ZIKV emergence and average results over 100 independent simulations. Other parameters are set to default values.

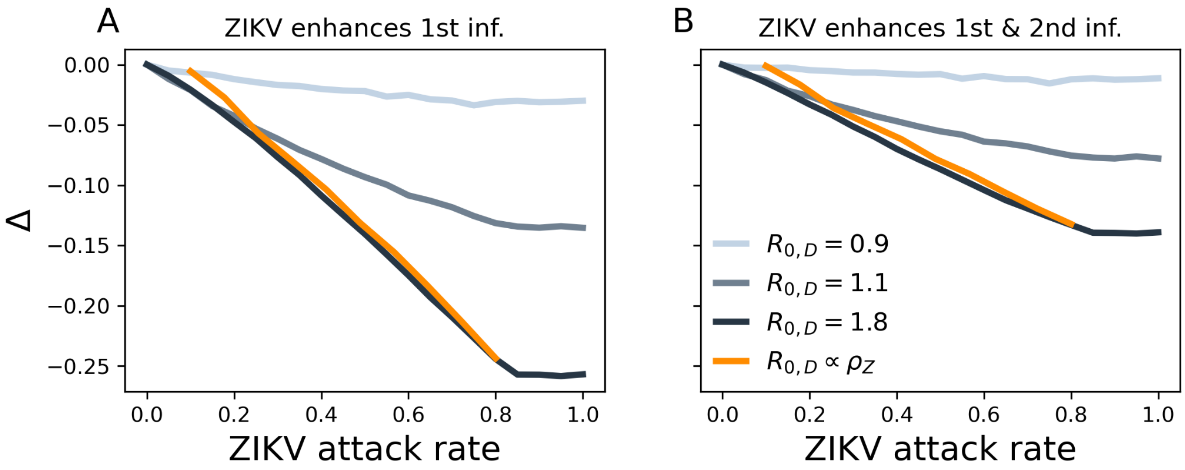


**Figure S15. Average age shift under different DENV transmission scenarios.** Panels show Δ as a function of ZIKV attack rate, under the assumptions that ZIKV enhances primary DENV infections only (A) or both primary and secondary infections (B). Incidence weights $w_n^{-/+}$ are the same as those specified in Fig. 4 in the main manuscript. Grey lines correspond to scenarios where $R_{0,D}$ is held constant as $\rho_Z$ is varied. In contrast, the orange line corresponds to a scenario where $R_{0,D}$ and $\rho_Z$ co-vary due to shared drivers of transmission. In particular, we a assume a linear relationship: $R_{0,D} = 1.29(\rho_Z - 0.1) + 0.9$, where $\rho_Z$ ranges from 0.1 to 0.8. In this scenario, values of Δ are comparable to those observed under high DENV circulation ($R_{0,D} = 1.8$). Of note, the relation between Δ and $\rho_Z$ is steeper in this case, meaning that co-variation in transmission can exacerbate the effect of ZIKV on Δ. In addition, the steeper slope implies that Δ hits 0 faster as $\rho_Z$ is decreased. Here, we let DENV spread for 100 years before introducing ZIKV and calculate Δ using incidence collected during the fourth year since ZIKV emergence. Other parameters are the same as in Fig. S6. Results are averaged over 300 simulations.
